# Thalamic connectivity-based Biomarkers for Neuromodulation in Patients with Refractory Epilepsies

**DOI:** 10.1101/2025.02.05.25320539

**Authors:** Giovanna Aiello, Daniel J. Soper, Peter N. Hadar, Angelique C. Paulk, Sydney S. Cash, Rafael Polania, Lukas Imbach, Pariya Salami

## Abstract

Thalamic electrical stimulation is being used as a treatment for drug-resistant epilepsy, yet current strategies benefit only about half the patients. Advancing patient outcomes necessitates tailored target selection for stimulation including the development of biomarkers for successful treatment. Here, we analyzed intracranial recordings of the most discussed thalamic nuclei for neuromodulation in epilepsy: anterior nucleus (ANT), centromedian nucleus (CM), pulvinar (PLV), and mediodorsal nucleus (MD) in 38 patients, to evaluate how their functional connectivity to epileptogenic areas may differ from that of non-epileptogenic areas in both sleep and wake states. We found that the thalamocortical connectivity to seizure onset areas is region and frequency-dependent. Additionally, we showed that thalamic connectivity is modulated in high versus low seizure risk conditions, further emphasizing its role in ictogenesis. Most importantly, we used the connectivity signature to predict the outcomes in patients receiving RNS within CM or PLV for neuromodulatory treatment and showed that these connectivity measures could serve as biomarkers of responsiveness to the treatment. These findings suggest that interictal connectivity data can inform patient-specific neuromodulation targeting, providing insights into the neurostimulation efficacy for specific epilepsy phenotypes.

## 1 Introduction

Epilepsy is a prevalent neurological disorder^1,2^ impacting about 1% of the global population. Drug-refractory epilepsy (DRE), where adequate response to antiseizure medications (ASMs) is lacking, affects approximately one-third of patients^3(p20),4^, and brain stimulation is increasingly considered as a valid treatment option for these patients^5–7^. The thalamus has extensive anatomical connectivity to different brain regions^8^ and is the most common target for neurostimulation in DRE. Even though thalamic stimulation through both deep brain stimulation (DBS)^9–14^ or responsive neurostimulation (RNS)^6,15–19^ holds promising results for patients with epilepsy, there is still substantial variability in responsiveness to therapy^20^. Median seizure reduction rates after 5 years for anterior thalamic DBS are reported at 65% in the SANTE trial^21^ and 56% in the MORE study^22^. In the ESTEL clinical trial (centromedian DBS), the median seizure reduction was 53.8%^23^. One factor contributing to these inconsistencies is the variability in seizure onset location. While DBS targeting the anterior nucleus of the thalamus (ANT) is shown to reduce seizure frequency up to 86% in frontotemporal epilepsies, this reduction only reaches 39% in patients with seizure onsets located elsewhere^14^. This variation in clinical efficacy might be related to our lack of knowledge in identifying optimally effective targets for stimulation for a specific patient. Indeed, the thalamus alone consists of approximately 60 subnuclei^24,25^, all with different anatomical and histological properties, and potentially different involvement in epilepsy^26–28^.

It has been now well-established that epilepsy is a network disorder^29–34^, and the thalamus, strongly connected to cortical and subcortical areas, is proposed to play a key role in this network^35^. Moreover, it is suggested that the connectivity of a given thalamic nucleus to a specific brain area reflects its involvement in the seizure activity emerging from that area^9,27^. Therefore, stimulation of a specific thalamic nucleus may differentially impact identified seizure networks^9^. Indeed, a growing number of studies aim at identifying the best target for thalamic neuromodulation, however these findings are not conclusive, and the best candidate nucleus for implantation for each epilepsy type has not been defined yet^9^. A better understanding of tailored implantation targets is required, but most studies either focus solely on temporal-lobe epilepsies^36,37^ or investigate a single nucleus^10,15,38–41^.

In this study, we hypothesized that by generating a comprehensive characterization of the functional connectivity profile of the primary thalamic nuclei targeted in epilepsy therapies (anterior (ANT), the centromedian (CM), the pulvinar (PLV) and the dorsomedial (MD) nuclei of the thalamus)^28^, we can identify their involvement in the epileptic network and ultimately predict which nucleus can serve as the best neurostimulation target for the treatment of distinct seizure types. As the thalamus plays a key role in sleep regulation ^42–50^, and epilepsy is influenced by sleep and circadian rhythms^42,43,51,52^, we separately considered sleep/wake stages. We further examined whether this variability in connectivity is frequency-dependent and assessed how these measures are modified when the seizure risk increases.

We evaluated the thalamocortical connectivity profile of 38 patients who received intracranial implants within at least one thalamic nucleus. We found that the connectivity patterns with cortical and subcortical regions differ from one nucleus to another, and more importantly, this connectivity changes when these regions are within the seizure onset areas. In addition, we found that the connectivity-based measures correlate with the daily risk of seizure occurrence. Further, we demonstrated that these measures could serve as biomarkers to predict whether a patient may benefit from thalamic RNS treatments. These novel insights highlight the translational potential of thalamic biomarkers for improving epilepsy treatment and understanding the mechanisms behind neurostimulation efficacy.

## 2 Materials and Methods

### 2.1 Patient Population

We analyzed data of 38 patients (21 males, age at implantation (mean±std) = 30.1 ± 15.7 years old) with DRE (age of onset (mean±std) = 14.7 ±15.6 years old), who were admitted to the Epilepsy Monitoring Unit (EMU) for presurgical evaluation to identify their epileptic foci (Table 1). Patients were admitted to the EMU of Massachusetts General Hospital (Boston, USA) or the Swiss Epilepsy Clinic (Zurich, Switzerland) between 2020 and 2024 for a minimum of 4 and a maximum of 15 days and monitored with intracranial stereo electroencephalography (sEEG) electrodes. The sEEG depth electrodes (PMT, Chanhassen, MN, USA) with diameter 0.8 and 4-16 platinum/iridium-contacts (electrodes) 1-2.4 mm long with inter-contact spacing ranging from 4-10 mm were placed stereotactically, based on clinical indications, for seizure localization. The electrode locations were determined by the clinical team independent of this study. Only patients who received at least one contact in one of the four thalamic nuclei of interest were selected for analysis. Seizures were reviewed by expert epileptologists so that the seizure onset zone (SOZ) and the time of onset were identified. The subjects’ consent was obtained according to the Declaration of Helsinki and was approved by the Institutional Review Board (IRB) at Mass General Brigham and the ethics committee of the Canton of Zurich (KEK). Following presurgical evaluation, patients received treatments such as resection or RNS (details in Table 1) based only on clinical decision making and without regard for this research. Patients who received Responsive Neurostimulation (RNS, Neuropace) in the PLV/CM nucleus (N=4 and N=8 respectively) were further followed up to determine biomarkers predicting their responses to RNS.

**Table 1.** Patient Demographics. Patient demographics (including gender, Engel scale P.I. if Neuromodulation, age of onset, age at implantation, treatment plan, nuclei) is reported. **Abbreviations: OA:** Onset age, **IA:** Implantation age, **RNS:** Responsive Neuromodulation, **ANT:** Anterior nucleus of the thalamus (AV), **CM:** Centromedian nucleus of the thalamus (CM), **PLV:** Pulvinar nucleus of the thalamus (PuM), **MD:** Mediodorsal nucleus of the thalamus (MDm), **LITT:** Laser Interstitial Thermal Therapy, **HC:** Hippocampus, **STG:** Superior Temporal Gyrus, **Front:** Frontal, **LT:** Lateral temporal, **MT:** Mesial temporal, **Par:** parietal, **Oc:** occipital, **Cing:** cingulate, **Subc:** subcortical, **Ins:** insula, **Thal:** thalamus, **MTS:** Mesial temporal sclerosis, **TSC:** tuberous sclerosis, Reported in brackets of the nuclei is the name of the selected nucleus by the FreeSurfer thalamic parcellation. In bold, patients who received CM or PLV RNS for which responsiveness to therapy is known. Engel class is reported.

| Patient / Gender / Engel | OA / IA | SOZ | Treatment choice | Thalamic nuclei |
| --- | --- | --- | --- | --- |
| **1/M IVB** | **5/31** | **Front (L) + LT (R) + Bi-Parietal** | **CM RNS (Non-responder)** | **CM** |
| 2/M | 1/10 | Front (R) + MT(L) | Resection | CM, MD |
| 3/M | 0/8 | Par (R) | Resection | CM, MD |
| **4/M IVB** | **6/9** | **LT (L) + Bi-Occipital + Bi-Parietal + Thal (R)** | **CM RNS (Non-responder)** | **CM, PLV** |
| **5/F IVC** | **1/21** | **Bi-Cingulate + Bi-Frontal + Subc (R) + Bi-Thalamic** | **CM RNS (Non-responder)** | **CM** |
| 6/M | 5/11 | Cing (L) + Bifrontal + Bisubcortical | CM RNS | MD |
| 7/M | 6/25 | MT (R) | Resection | CM |
| 8/F | 1/16 | LT (R) + MT (R) + Oc (R) + Subc (R) | Resection | CM |
| **9/F IVB** | **0/24** | **LT (L)** | **Resection + VNS + CM RNS (Non-responder)** | **CM** |
| **10/F IIIA** | **13/17** | **LT (R) + Bi-MT** | **PLV RNS (Responder)** | **ANT, PLV** |
| **11/F IIIA** | **6/34** | **Front (R) + LT (R) + Bi-MT** | **Resection + PLV RNS (Responder)** | **ANT, MD, PLV** |
| 12/M | 62/66 | MT (L) | Resection | ANT |
| 13/M | 3/12 | Insula (L) + LT (L) + MT (L) + Par (L) + Subc (L) | None | ANT, CM, MD, PLV |
| **14/M IVA** | **6/29** | **Bi-Cingulate + LT (L)** | **Resection + CM RNS (Non-Responder)** | **CM** |
| 15/F | 26/41 | Bi-frontal + MT (R) | Resection + HC RNS | ANT, MD, PLV |
| 16/F | 1/37 | Bi-LT + Bi-MT + Oc (L) | Resection | CM |
| **17/F IIIA** | **7/15** | **MT (R) + Bi-Parietal + Bi-Thalamus** | **CM RNS (Responder)** | **CM, MD** |
| **18/F IIB** | **9/17** | **Bi-Frontal + Subc (R)** | **CM RNS (Responder)** | **CM** |
| 19/F | 2/23 | Ins (R) + Bi-MT + Subc (R) | LITT | CM |
| 20/F | 8/30 | Subc (L) | LITT+Resection | ANT, CM, MD |
| 21/F | 9/16 | Cing (L) + Bi-MT + Oc (L) | LITT | ANT |
| 22/M | 39/59 | MT (L) | Resection | CM, MD |
| 23/M | 18/24 | Front (R) + Subc (R) | LITT | CM, MD |
| **24/M IB** | **11/32** | **Bi-Frontal + LT (L) + MT (L) + Par(R)** | **CM RNS (Responder)** | **ANT, CM** |
| 25/M | 19/31 | Cing (R) | LITT | CM, PLV |
| 26/M | 19/26 | Bi-MT | HC RNS | ANT, PLV |
| 27/F | 3/21 | Par (L) | Resection + Frontal RNS | CM, MD, PLV |
| 28/M | 6/22 | MT (R) | Resection | PLV |
| **29/M IVB** | **33/36** | **LT (L) + Oc (L) + Par (L)** | **PLV RNS (Non-Responder)** | **PLV** |
| 30/M | 19/28 | Bi-Cingulate | LITT | ANT, CM, MD |
| 31/F | 25/38 | Bi-Frontal + MT (R) | Resection | CM |
| 32/M | 6/54 | LT (L) + MT (L) + Oc (L) | PLV RNS | PLV |
| **33/F IVA** | **0/16** | **Front (L) + LT (L) + Oc(L)** | **STG/PLV RNS (Non-Responder)** | **CM, PLV** |
| 34/F | 52/57 | Front (R) + Thal (R) | None, planned RNS | CM, MD |
| 35/M | 40/55 | Cing (R) + Front (R) + MT (R) | Resection | ANT, CM, PLV |
| 36/F | 29/53 | Bi-MT | None | ANT, PLV |
| 37/M | 32/53 | Bi-MT + Thal (L) | HC RNS | ANT |
| 38/M | 32/45 | Par (L) + Cing (L) | Thermocoagulation + Planned Resection | ANT |

### 2.2 Electrode Implantation and Localization

We determined the anatomical position of each contact through a combined approach involving volumetric and surface registration. Electrode coordinates were manually extracted from CT scans and placed within the native space of each patient. Subsequently, FreeSurfer was utilized to identify brain regions, and an electrode labeling algorithm (ELA) was employed to map electrodes to the Desikan-Killiany-Tourville (DKT) atlas^53–59^. For patient 38 (from the Swiss Epilepsy Clinic), the electrode location was verified by direct localization in post-operative MRI using Sure-Tune and LeadDBS^60,61^. Channels were categorized into eight groups for both the left and right hemispheres based on proximity and structural similarity:

1. Mesial temporal structures (including hippocampus, amygdala, subiculum) [referred to as MT]
2. Lateral temporal areas (encompassing inferior, middle and superior temporal) [referred to as LT]
3. Centroparietal areas (covering pre-central, post-central, superior and inferior parietal) [referred to as Par]
4. Frontal areas (encompassing frontal areas, orbitofrontal) [referred to as Front]
5. Occipital structures (including occipital area, cuneus, and lingual) [referred to as Oc]
6. Subcortical structures (including nucleus accumbens, substancia nigra, basal ganglia, caudate, putamen, cerebellum, choroid plexus, claustrum, pallidum) [referred to as SC]
7. Insula [referred to as Ins]
8. Cingulate [referred to as Cing]

To localize electrode contacts within the thalamus, we utilized the electrode volume labeling (EVL) approach ^53^. The segmentation of the thalamus was carried out following the labeling of brain regions, including the brain and subcortical structures, using FreeSurfer ^62^. The outcomes of thalamic segmentation were exported as volumes and transferred to MATLAB (MATLAB 2020b), where enclosed volumes were created for each brain region label^53^. In reference to the initial thalamic nucleus segmentation in FreeSurfer, the four target nuclei were identified as follows: CM (Centromedian), AV (Anterior), PuM (Pulvinar) and MDm (Dorsomedial).

### 2.3 Data Acquisition and Processing

After patients were admitted in the EMU, they were recorded continuously for 24-hours a day and multiple days. Throughout each day, data was classified into sleep and wake periods using SleepSEEG^63^ for sleep staging, to finally select 15-minute segments of NREM (N2, N3 stages), REM, and AWAKE. NREM periods were kept if at least 60% of their 30-second epochs were selected as such, else they were rejected from the analysis. REM sleep was excluded due to its seizure-suppressing nature^64^ and low representation in our data (Supplementary Figure 1D). All data were interictal, collected at least 30 minutes away from seizures. Data were visually inspected and the parts that were contaminated with artifacts (e.g., signal saturation or movement) were excluded from the analysis. Recordings were made with Natus Quantum (Natus Medical, Pleasanton, CA) at a 1024 Hz sampling rate. The data analysis was performed using **MATLABR2023b** (using the open-access toolbox *Fieldtrip*^65^) and in **Python** (using the open-access toolbox *MNE* ^66^). Data were re-referenced to a bipolar montage and bandpassed from 1-100 Hz, Notch-filtered for line noise. Data were divided into 4-second non-overlapping windows, and for each window spectral-domain features were computed using a multi-taper Fourier transform between 1-90 Hz, with data zero-padded to the next power of 2 and a ±3 Hz smoothing window (frequency resolution: 0.25 Hz). Power spectra were normalized by total power, and frequency bands were defined as *δ* (1-4 Hz), *θ* (4-8 Hz), *α* (8-12 Hz), *β*_1_ (12-20 Hz), *β*_2_ (20-30 Hz), *γ*_1_ (30-60 Hz), *γ*_2_ (60-90 Hz). ^65^.

### 2.4 Feature Extraction

For each patient, recording electrodes were classified into two groups: 1) those located within the seizure onset zone (SOZ) and 2) those located in non-Epileptogenic (NE) areas. The Seizure Onset Zone (SOZ) was visually identified by expert epileptologists as the channels where the seizure initiates. Contacts that were located close to - but not definitively inside - the seizure onset areas and showed fields of activity at the seizure onset were removed from the analysis. We then assigned all the contacts to their respective region according to the brain region localization procedures (see above).

Hereafter, when referring to SOZ vs NE comparisons, we will be focusing on those contacts that belong to SOZ and those that belong to NE within the same region (for instance, mesial temporal). Unless specified, values of the extracted features were averaged among contacts belonging to SOZ and NE for the same region, leading to one value for each category.

#### 2.4.1 Connectivity Measures

The connectivity of each nucleus of interest with the regions within the same side (ipsilateral) was computed using the weighted phase-lag index debiased (*wPLId*)^67^. *wPLID* measures the degree of consistency of phase lag between two signals across time intervals (epochs) and is our preferred measure of connectivity since it is robust to volume conduction (zero-phase lag), sample size, and signal amplitude. The recordings were divided into 4-second epochs with no overlap. We computed *wPLId* between the contact within the thalamic nucleus of interest and every other contact throughout the ipsilateral brain hemisphere, obtaining a matrix of dimensions 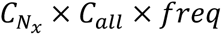, where 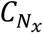 are the contacts in the nucleus of interest, *C*_*all*_ are the contacts within other areas in the rest of the ipsilateral brain and *freq* is the frequency array (1 to 90 Hz). We then computed the average connectivity values in each band of interest.

When computing connectivity, we focused on the *differential* connectivity from each thalamic nucleus *N*_*x*_ to SOZ and to NE within the same region *i,* which we define here as the Connectivity Divergence Index (CDI):

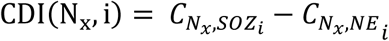

The CDI was always computed within-subject to avoid inter-subject variability and to minimize the data imbalance. We ran paired (Wilcoxon signed-rank) tests to understand potential differences between SOZ and NE areas. The p-values were corrected by dividing by the number of bands to identify which combinations of band-regions were significantly different between SOZ and NE areas.

#### 2.4.2 Epileptogenic-Non epileptogenic Index

To explore whether the connectivity of the nucleus of interest with the rest of the brain is frequency dependent and whether this connectivity varies from SOZ to NE areas for each band, we computed the following measures:

1. Epileptogenic frequency occurrence (EFO), indicating how often (across samples) the connectivity between the thalamic nuclei and epileptogenic areas (seizure onset zones) peaks within a specific band (defined as *foi*, frequency of interest).
2. Non-epileptogenic frequency occurrence (NEFO), indicating how often the connectivity between the thalamic nuclei and non-epileptic areas peaks within a specific band.

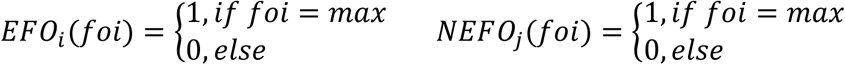

These values were computed for each frequency and were further averaged across regions within each network:

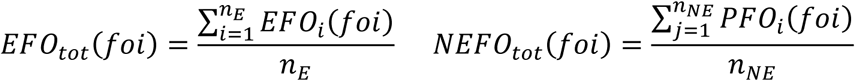

with *n*_*E*_ indicating the number of epileptogenic network samples, and *n*_*NE*_ indicating the number of non-epileptogenic network samples.

Finally, we computed the Epileptogenic-Non epileptogenic Index (ENI), a metric we define as the ratio of EFO to NEFO for each frequency band:

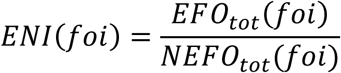

An ENI>1 for a given frequency band indicates that the connectivity within that band was more prominent in the epileptogenic network (seizure onset zone; SOZ) than in the non-epileptogenic network.

For each nucleus, we computed the ENI metric separately for each frequency band and region. This resulted in a multidimensional (1 × *Regions*) vector of ENI values, which we represented as a polygon with *N*_*Regions*_ vertices. The area of this polygon was calculated: the greater the area, the more the frequency band accounts for epileptogenicity in that nucleus (intended as, pathological activity related to a seizure-inducing network).

To test whether ENI was significantly higher than 1 (i.e., the peak of connectivity was more often in that frequency band for epileptogenic than non-epileptogenic networks), we ran a one-sided *χ*^2^ test.

We ran the same algorithm within each region separately. For each region and each nucleus, we considered the polygon of pathogenicity in each frequency band (i.e., the polygon had *N*_*FrequencyBands*_ vertices). In this case, the greater the area, the more that nucleus is associated with epileptogenicity in that region.

### 2.5 Correlation to Seizure Occurrence

To determine whether the changes in thalamocortical connectivity correlate with the likelihood of seizure occurrence for each selected interictal period, we measured the following values:

a. Number of seizures in a 24-h window (±12ℎ), centered on the period of interest. Here after, “*Count*”.
b. Time to the next seizure (minutes). Here after, “*T*_*next*_”.
c. Time to the previous seizure (minutes). Here after, “*T*_*previous*_”.

We then defined the *Epileptogenicity Index* of each analyzed period as the following:

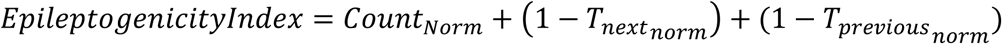

where the times and seizure counts were normalized for each patient according to their maximum value. We then ran linear mixed models (LMM) to assess whether the Epileptogenicity Index of the recorded period was correlated significantly to any of the connectivity or power spectrum measures, using the patient as a Random Effect, according to the following model:

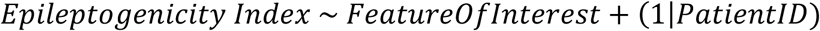

The normalization itself removes differences in the range of the Epileptogenicity Index, while the random effect removes inter-subjects’ variability in the feature of interest and thus provides a more robust measure than simple linear regression models.

From the LMM, we consider the t-statistics of the fixed effect only if it was statistically significant. We ran the model separately for each band (in Power Spectrum) and each band-region combination (in Connectivity). We separately computed our measures of interest on days with a seizure (i.e., *Count* ≥ 1) and seizure-free days (i.e., *Count* = 0).

### 2.6 Responsiveness to RNS

Out of the patients who received CM thalamic implants during their presurgical evaluation, N=8 patients received CM RNS, and N=4 received PLV RNS. We divided patients into responders (Engel I-III) and non-responders (Engel IV), based on the changes in their seizure frequency after receiving RNS stimulation. We assessed whether the connectivity and power spectrum measures from the data recorded during the presurgical evaluation could predict patients’ responsiveness to therapy.

### 2.7 Statistical Analysis

Due to the intrinsic non-normality of our data, we have only considered non-parametric tests. To compare nuclei connectivity to different brain regions, we ran multivariate ANOVA with post-hoc Tukey-HSD multiple comparisons correction. To evaluate whether CDI was positive or negative for each nucleus-region pair, or differences within-subjects (such as CDI in AWAKE vs NREM), we ran paired Wilcoxon signed-rank tests, accounting for multiple comparisons by Bonferroni correction. Our ENI measure was tested for significance relative to zero by running a *χ*^2^ test. In the analysis where two independent groups have been tested for differences (such as the responsiveness analysis), we used the Mann-Whitney rank-sum test, accounting for multiple comparisons by Bonferroni correction. The correlation analysis was run using Linear Mixed Models, which account for inter-subjects’ variability and remove single-patient influence. The results are reported as follows: * (p<0.05), ** (p<0.01), *** (p<0.001), **** (p<0.0001). Further details on which test was deployed are reported in the Results section.

### 2.8 Data availability

All the results needed to evaluate the conclusions in the paper are present in the paper and/or the Supplementary Materials. The raw data of this study are available upon request from the corresponding author.

## 3 Results

### 3.1 Study design

We analyzed intracranial data from N=38 patients, who received stereo electroencephalography (sEEG) electrode implantation in at least one of the four nuclei of interest during their multi-day presurgical evaluation (Figure 1A,B). Figure 1 Contacts targeting cortical and subcortical areas were grouped based on proximity and structural similarity (see Methods; Figure 1D). To assess if experimental conditions, such as the number of recording days, number of implanted contacts, number of seizures, and sampled regions within the brain were comparable, we analyzed their distribution across the four sub-cohorts. ANT-implanted patients had leads mostly in the frontotemporal, PLV-implanted patients in the temporo-posterior, MD in the frontotemporal/parietal and CM-implanted in spread areas. This is attributable to clinical reasons, as the implantation plan was independent of this research study. There were no significant differences in the days the patients were recorded or the number of electrodes they received (Kruskal-Wallis, p>0.05). We found differences in the number of seizures per patient (Kruskal-Wallis, p<0.05), with MD and CM -implanted patients presenting a higher number of seizures. For detailed results, see Supplementary Figure 1.

**Figure 1:**
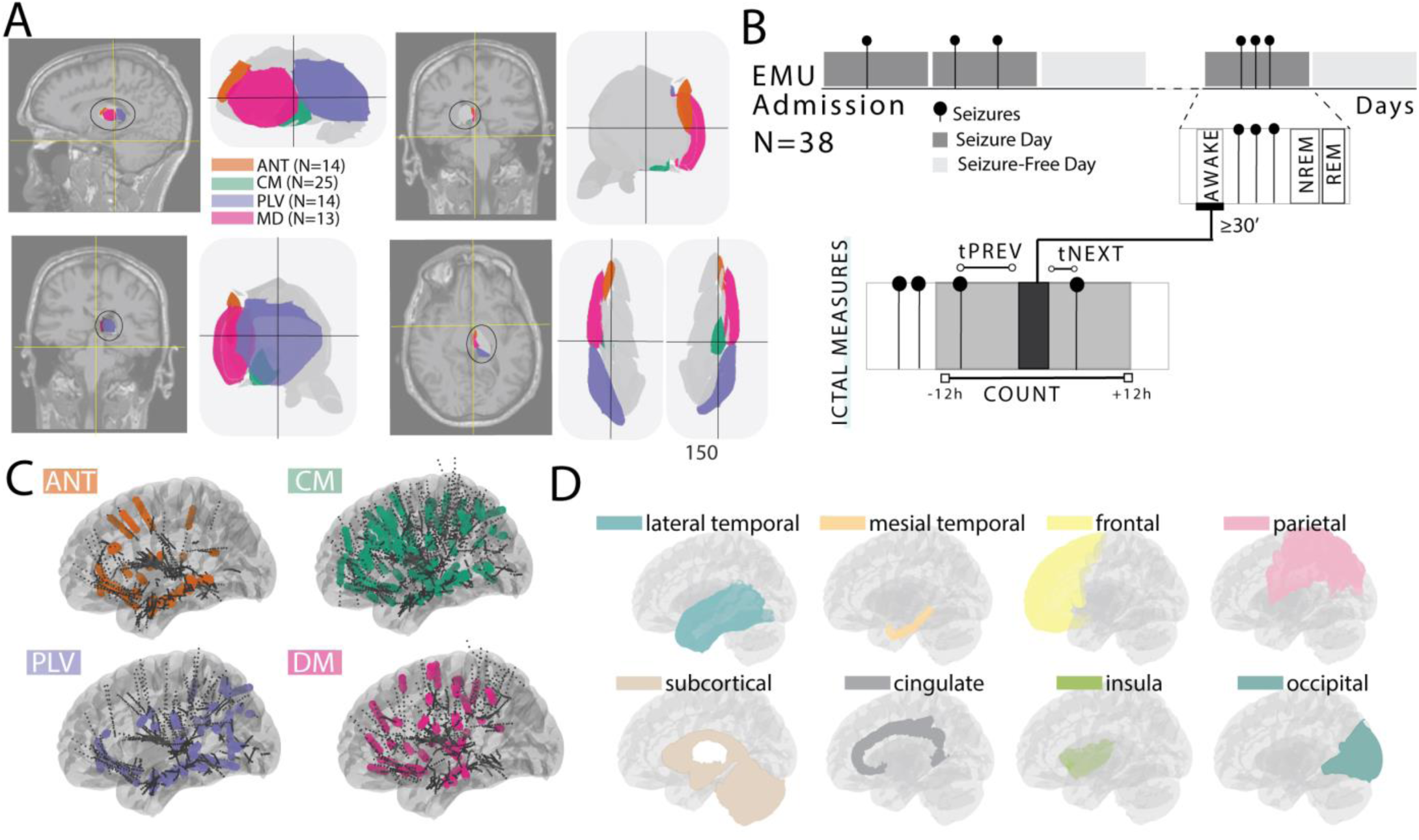
Study Design. **A)** Sagittal, Coronal (Anterior and Posterior) and Horizontal view of the thalamic nuclei and their placement. **B)** Schema of data selection: after admission to the EMU, patients are recorded for consecutive days. Interictal periods (15 min from each day) from AWAKE, NREM, and REM periods, at least 30 minutes away from the seizure were selected. tPREV = time to the previous seizure, tNEXT = time to the next seizure, COUNT = amount of seizures **C)** Number of sEEG/ECoG contacts for each subcohort (count density). **D)** Regions definition.

### 3.2 Connectivity profiles vary across thalamic nuclei

The functional connectivity of the four thalamic nuclei with the rest of the brain was calculated using the weighted phase-lag index debiased (*wPLId*) metric. We found that ANT is predominantly connected to mesial temporal regions, with statistically higher connectivity than MD and CM (p<0.001, ANOVA-N), but the ANT connectivity to mesial temporal areas was not statistically different from that of PLV (Figure 2A-B). CM and MD showed overall similar connectivity to different areas, but CM was more strongly connected to parietal areas and MD to mesial temporal areas (p<0.001) (Figure 2A-B). CM connectivity to parietal areas was significantly stronger than those of all other nuclei (MD, ANT and PLV; p<0.001). PLV, meanwhile, was mainly connected to lateral temporal and occipital regions but also had significant connections to mesial temporal areas (Figure 2A-B). ANT and PLV had significantly lower broad-band average connectivity than CM and MD (Figure 2A, inlet, p<0.0001).

**Figure 2:**
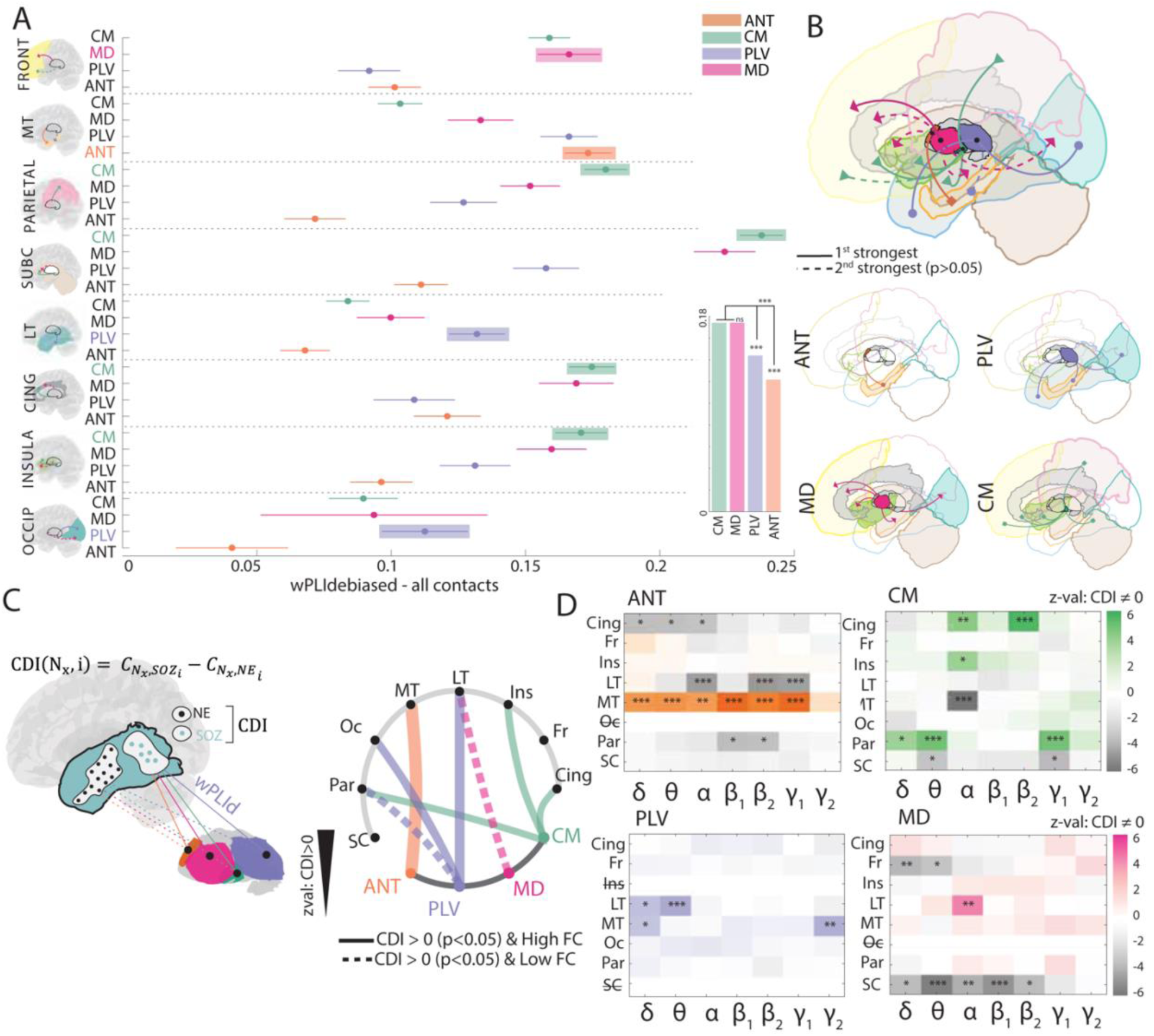
Nuclei-based profiles of Connectivity and Epileptogenic Networks Specificity. **A)** Connectivity distribution for each sub-region and nucleus as reported by ANOVA-N with post-hoc Tukey-HSD (*μ* ± 95% *CI*). Connectivity is reported as broadband connectivity across all contacts (SOZ and NE). When CI intervals between two nuclei overlap, their difference is not statistically significant. For each region, the highest-connected nucleus (preferential) is highlighted, and the two highest connections are reported in the brain representation. In the inlet, Average broadband, spread wPLI (from Figure 2A) is reported for each nucleus, with statistical significance between nuclei (post-hoc Tukey HSD) (left). **B)** For every region, 1^st^ (solid) and 2^nd^ (dashed, not significantly different from 1^st^) preferential connections with the nuclei are reported (top). The same result is reported for every nucleus separately (bottom). **C)** For each nucleus and region, we computed the CDI as the difference in nucleus connectivity to SOZ and NE. We then tested for differences using Wilcoxon signed-rank test. We report one-tailed significant comparisons (where CDI∼0, and z-val>0). The thickness of each line in the chordplot is proportional to the z-value. If the pairing was also presenting high functional connectivity, it is represented with a solid line. Else, it is represented with a dashed line. **D)** Two tailed results from the Wilcoxon signed-rank tests are reported separately for each combination of band/region. Results have been corrected for the number of bands and regions.

We ran the same analysis purely considering nuclei connectivity to different regions only for SOZ contacts. We found mirroring results, where the strongest-connected nucleus to each region remained consistent, except for frontal (where CM prevailed) and insula (where ANT prevailed) regions (Supplementary Figure 2A-B). Interestingly, we found that broad-band ipsilateral connectivity increased for ANT, CM, and MD nuclei significantly in NREM states, while the PLV did not show any meaningful changes (Supplementary Figure 2D). When looking at the interaction of region and band specific effects, we noticed a consistent increase in connectivity in *α*, *β*_1_ band connectivity in NREM for the four nuclei and all regions (Supplementary Figure 2E). This was similarly reflected in NE and SOZ areas, although, for SOZ, we could not find statistically significant results, which we believe is to be attributable to sample size (Supplementary Figure 2F).

Functional connectivity is influenced by baseline differences in regional connectivity; hence we explored how the newly introduced Connectivity Divergence Index (CDI) metric, defined as the difference between nucleus-SOZ connectivity and nucleus-NE connectivity within the same region, could provide further insights into epilepsy-specific network connectivity. Figure 2C illustrates CDI values identifying greater connectivity to SOZ contacts versus NE contacts across nuclei and regions, corresponding to a significantly positive CDI (one-tailed Wilcoxon signed-rank test). Some previously identified high connectivity combinations were confirmed (ANT-MT, PLV-Occ, PLV-LT, CM-Ins, CM-Cing, CM-Par), while new significant connections with positive CDI were observed, such as MD-LT and PLV-Par, suggesting additional roles of those nuclei in epileptogenic networks arising from these regions. The same result is shown separately for NREM and AWAKE in Supplementary Figure 2C.

To determine the relevance of positive and negative (i.e., selectively higher connectivity to non-seizure areas) CDI values, we analyzed all significant CDI combinations (CDI ≠ 0, two-tailed Wilcoxon signed-rank test, Bonferroni correction) in Figure 2D (methodology is shown in Supplementary Figure 2G). We report significant negative CDI for ANT-LT, ANT-Cing, ANT-Par, CM-LT, CM-Subcortical, MD-Subcortical and MD-Front. Interestingly, while some regions consistently show positive or negative CDI regardless of the frequency band (for instance, ANT-MT), others had a band-specific CDI, indicating frequency-dependent connectivity values.

Patients with a SOZ in a specific region (for example, in the *frontal* area) could have some electrode contacts in specific locations that are non-epileptogenic (NE) yet still belong to the frontal area. In this case, the relative NE contacts have been named NE+. On the contrary, patients who do *not* have a SOZ in the frontal region, but still have NE contacts in frontal locations, were defined as NE-. As a control, for every region, we analyzed differences between NE+ and NE-, across patients with unpaired Mann-Whitney tests. We found that CDI values previously reported were due to within-subject specific differences in SOZ and NE areas, as the same regions did not show consistent significance when analyzing NE+ vs. NE-.

We also investigated differences between SOZ connectivity in patients whose SOZ was in that region (SOZ+) vs NE connectivity in patients whose SOZ was not in that region (NE-) with unpaired Mann-Whitney tests. While this test was run across subjects, testing for unpaired differences in SOZ+ vs NE showed similar results as in Figure 2E (Supplementary Figure 2I).

### 3.3 Epileptogenic networks are characterized by specific frequency band signatures

Having established that specific nuclei are differentially associated with epileptogenic or non-epileptogenic networks, we aimed to determine if this variability in connectivity is frequency-dependent (Figure 3A).

**Figure 3:**
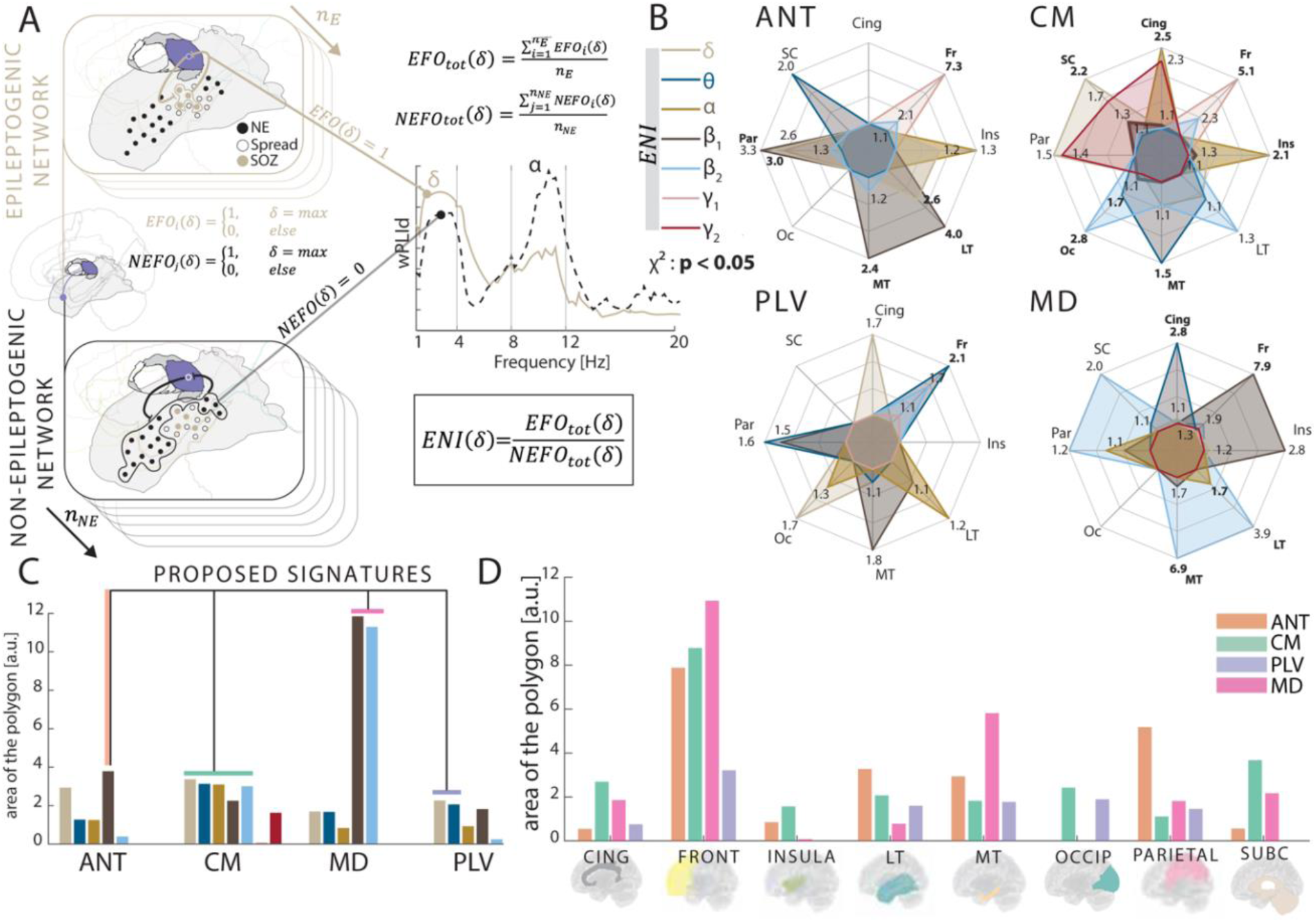
Nuclei-specific frequency biomarkers of epileptogenic networks. **A)** We separated each nucleus-region connectivity into epileptogenic (i.e., nucleus-SOZ) and non-epileptogenic (i.e., nucleus-NE) networks. By examining the connectivity spectrum, we determined when each frequency band presented as maximum and counted the number of times for which its connectivity was greatest across all bands. We then determined the ENI as the ratio of occurrences between epileptogenic and non-epileptogenic networks (EFO and NEFO) for each band. **B)** For every nucleus and every band (encoded as a color), we report the ENI at every region. If significant (*χ*^2^ test), the region and the relative ENI is in bold. The band with the highest area is represented with a thick line of the corresponding polygon. **C)** Based on the plots in B, we report the area of the polygon for each nucleus for each frequency band, highlighting for each nucleus the one with the highest area, which we propose as epileptogenic signature. **D)** We report the area for each nucleus in the analyzed regions.

For every nucleus-region pairing, we identified the Epileptogenic-Non epileptogenic Index (ENI), indicating the prevalent band in epileptogenic networks, and its statistical significance in each specific frequency band of interest. Intriguingly, we found that each nucleus behaved differently in terms of which band maximally separated the epileptogenic (seizure onset zones; SOZ) and non-epileptogenic areas (Figure 3B).

ANT-SOZ networks were mainly characterized by *β*_1_ band connectivity, specifically in mesial temporal (ENI = 2.4), lateral temporal (ENI = 4.0), and parietal regions. CM connectivity to SOZs peaked similarly in *δ*, *θ*, *α*, *β*_1_, *β*_2_ in several regions. Possibly due to its broad-band enhanced connectivity to SOZ, a wide range of regions presented significant differences to physiological networks, such as subcortical (ENI = 2.2), cingulate (ENI = 2.5), frontal (ENI = 5.1), insula (ENI = 2.1), mesial temporal (ENI = 1.5) and occipital (ENI = 2.8) regions. MD networks had an increased pathogenicity in *β*_1_, *β*_2_ bands, specifically in the frontal (ENI = 7.9), insula (*β*_1_) and subcortical/parietal/mesial temporal (ENI = 6.9)/LT (ENI = 3.9) networks (*β*_2_). Finally, PLV-SOZ networks were characterized by *δ*, *θ* bands, specifically for occipital, cingulate, parietal, and frontal (ENI = 3.4) regions. The reported ENI values are all statistically significant (*χ*^2^, p<0.05).

With the grounding hypothesis that a higher prevalence of certain frequency bands in epileptogenic vs non-epileptogenic network connectivity suggests those specific frequency band’s engagement within seizure onset areas, the following bands may be ictogenic signatures of the four nuclei: *β*_1_ for ANT, broad-band (*δ* − *β*_2_) for CM, *β*_1−2_ for MD, *δ* − *θ* for PLV (Figure 3C). ENI values for each band can be found in Supplementary Figure 3A.

We further computed the joint ENI area (across bands) calculated from polygons in Supplementary Figure 3B. We found that the following nuclei-region pairs have the largest difference between SOZ and NE networks: CM with cingulate, insula, and subcortical areas; ANT with mesial temporal, lateral temporal, and parietal regions; PLV with occipital areas; and MD with mesial temporal, and frontal areas (Figure 3D).

### 3.4 Interictal thalamic activity correlates to seizure risk

We also investigated the relationship between network measures and seizure risk to determine whether the changes in the connectivity of each nucleus and the rest of the brain correlate with the seizure occurrence. Unexpectedly, both SOZ and NE spectral power differed between seizure-free days and seizure days, with significant changes in frequency bands from *δ* (where power spectra increased on seizure days) to *β*_1_ (where power spectra decreased on seizure days) (Figure 4A). Building on these findings, we investigated whether thalamic power spectra also varied with seizure likelihood. Analysis of seizure-free vs seizure-days in thalamic power spectra are reported in Supplementary Figure 4A, together with correlation between SOZ power spectrum and nucleus power spectrum (Supplementary Figure 4E). We observed a positive correlation between thalamic power spectra and seizure risk in the lower frequency bands (*δ* − *θ*) and a negative correlation in the higher bands (*α* − *γ*_1_). This pattern held true across all nuclei, indicating a shared underlying mechanism (Figure 4B). The same result without the statistical mask (i.e., also showing non-significant correlations) is reported in Supplementary Figure 4F. The correlation between thalamic power spectrum and seizure risk is separately analyzed for the three measures (i.e., time to next seizure, time to previous seizure and count of seizures) in Supplementary Figure 4B.

**Figure 4:**
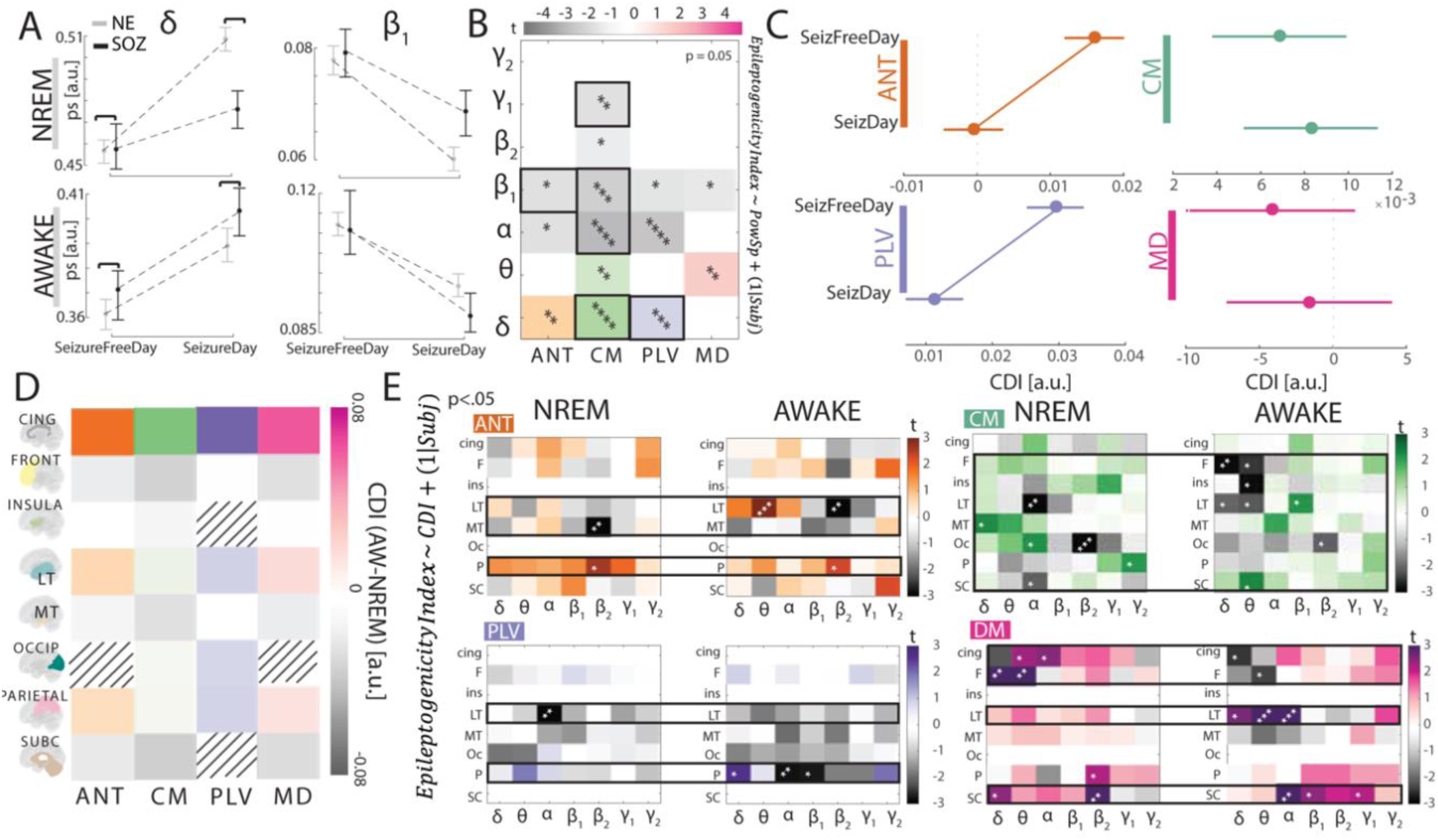
Thalamic activity and connectivity is state-dependent and correlates to seizure risk. **A)** Significant differences between seizure-free days and seizure-days identified by the strength of *δ*, *β*_1_. in SOZ and NE areas during NREM and AWAKE Dashed lines (seizure-free vs seizure day) and significance bars between SOZ and NE indicate statistical significance (respectively, Mann-Whitney rank-sum and Wilcoxon signed-rank tests, p<0.05). **B)** Results of Linear Mixed Models for prediction of the Epileptogenicity Index extracted from the strength of different frequency bands in all four nuclei. Results are reported as t-values, only if significant (p<0.05). **C)** CDI (Differential connectivity between SOZ and NE) in seizure-free and seizure days for each nucleus (*μ* ± 95% *CI*). **D)** Differences of CDI between AWAKE and NREM for all pairs of nuclei-regions. **E)** Results of Linear Mixed Models showing the regression of the Epileptogenicity Index to region-nucleus specific CDI. Results are reported as t-values, and statistical significance is indicated. Solid black boxes indicate statistically significance.

Next, we examined the effect of relative connectivity between SOZ and NE (using the CDI measure) on seizure distribution across different states. Interestingly, we observed that the CDI decreased on seizure days for ANT and PLV (p<0.001), while it did not change for the CM and MD nucleus (Figure 4C). For both ANT and PLV nuclei, on a seizure-day, the nucleus-NE connectivity was increased while nucleus-SOZ connectivity did not change compared to a seizure-free day (Supplementary Figure 4G), resulting in a decreased CDI measure. CM and MD, on the other hand, did not show significant changes at the group-level (Supplementary Figure 4G).

To further understand CDI’s interpretation and its modulation upon different brain states, we measured its change between sleep and wake states. Our analysis revealed that cingulate, lateral temporal, occipital, and parietal regions had a higher CDI in awake than in sleep states, while the opposite was true for frontal, insula, mesial temporal, and subcortical regions. While these trends were visible independently on the nucleus considered, p-values were not significant (p>0.05) (Figure 4D).

CDI modulates with both seizure occurrence and sleep/wake states. We further evaluated the relationship between CDI and the Epileptogenicity Index (defined in the Methods) (Figure 4E): specifically, CDI was able to predict the likelihood of seizure occurrence in extremely specific band-region pairs. The significant pairings (identified from the LMM) included mesial temporal, lateral temporal, and parietal for ANT; frontal, insula, lateral temporal, mesial temporal, occipital, parietal, and subcortical areas for CM; lateral temporal and parietal for PLV; cingulate, frontal, lateral temporal, and subcortical for MD. Of note, we had both positive and negative correlations: in certain specific bands/regions, CDI was negatively correlated to the Epileptogenicity Index (i.e., the higher the CDI, the lower the risk of a seizure); while in other cases, it was positively correlated (i.e., the higher the CDI, the higher the risk of a seizure). The correlation between the CDI and the Epileptogenicity Index was further analyzed separately for the three measures (time to next, time to previous, and number of seizures), yielding consistent results as to which nucleus-region combinations were relevant in seizure risk prediction (Supplementary Figure 4D). Correlations between CDI and time to the next seizure are separately shown in Supplementary Figure 5.

### 3.5 CM and PLV neurophysiological characteristics correlate to Responsiveness to RNS

The validity of measures such as the CDI depend on their correlation with clinical outcomes. Although not all patients received neuromodulatory interventions, we evaluated the relationship between connectivity and patients’ responsiveness to thalamic RNS in a small cohort of thalamic RNS implanted patients (CM-RNS: three responders, five non-responders; PLV-RNS: two responders, two non-responders). The patients’ seizure onset locations and Engel classifications are shown in Figure 5A1-A2 (more details in Table 1).

**Figure 5:**
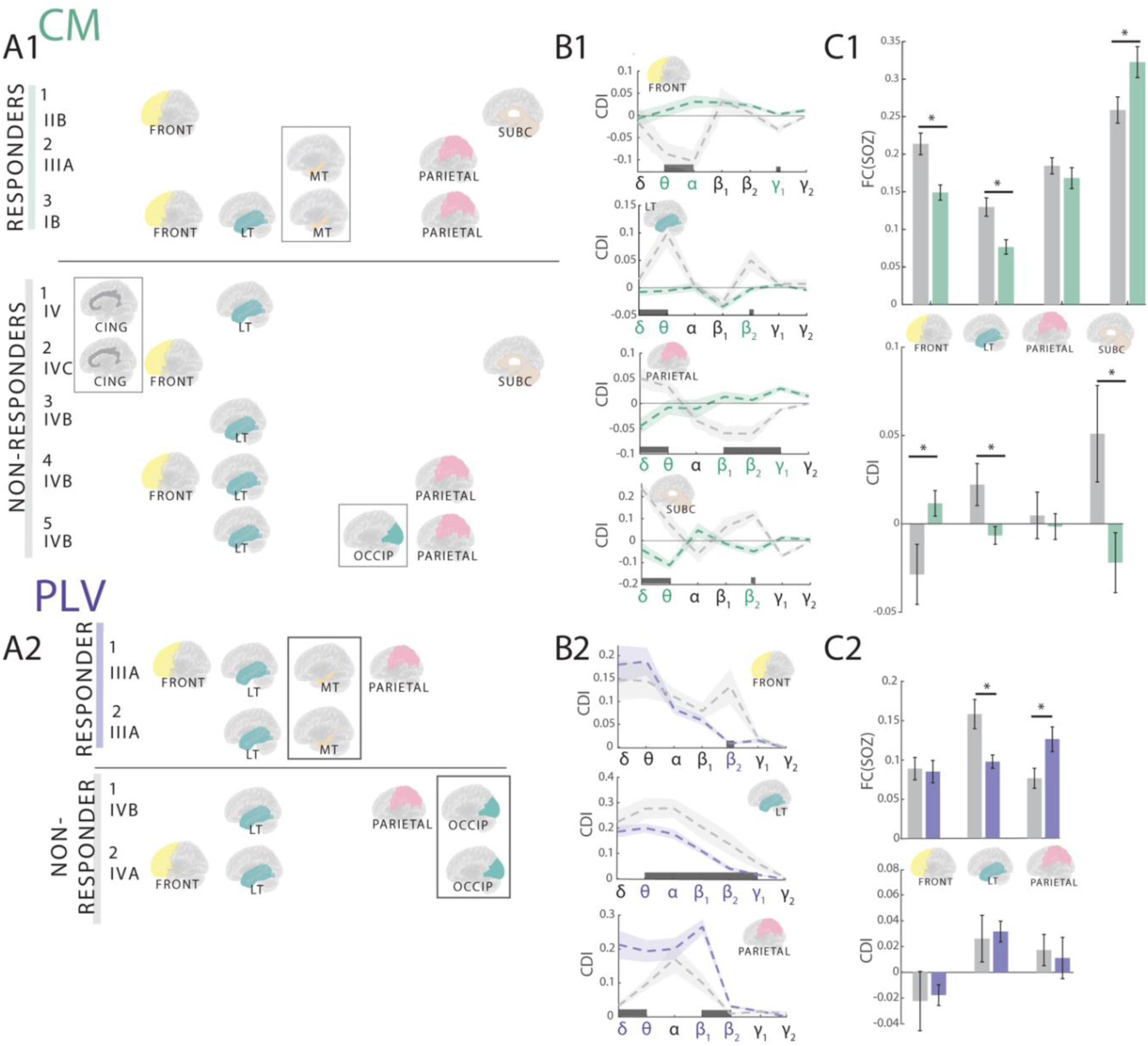
Thalamic-based biomarkers of responsiveness. **A1-A2)** Representation of seizure onset areas in patients who received RNS implants within CM (responders and non-responders with their respective Engel outcomes post-implantation). The grey boxes indicate the regions that could not be analyzed as only belonging to one group of responders or non-responders. **B1-B2)** CDI band-wise for each region in responders vs non-responders. The thick gray bars on X-axis indicate frequency bands for which the CDI difference between responders and non-responders was significant. **C1-C2)** Functional connectivity and CDI in responders and non-responders for each SOZ region, together with reported statistical significance (ANOVA-N with post-hoc Tukey-HSD).

Considering patients with CM-RNS, we found that non-responders had higher functional connectivity between CM and SOZ than responders in frontal and lateral temporal areas, while the opposite was true for subcortical regions (ANOVA-N with Tukey-HSD, p<0.0001) (Figure 5C1). Further analysis revealed that CDI was lower in responders compared to non-responders within lateral temporal and subcortical regions, while the opposite was true for frontal areas (Figure 5C1). No significant difference was found for connectivity to parietal areas. Interestingly, these differences were band-dependent (Figure 5B1). For all regions (except for the frontal), the key frequency bands differentiating between the two groups were *δ*, *θ*, and *β*_2_.

The same analysis was run on the PLV-implanted cohort, where we noticed a higher PLV-SOZ connectivity to lateral temporal (p<0.05) regions in non-responders compared to responders, with the opposite holding true for parietal areas (p<0.05), shown in **Figure 5B2**.

Comparing the CDIs, responders tend to have a higher CDI than non-responders in all regions except in parietal areas, although this was not significant (**Figure 5C2**). When looking at band-wise differences, we observed that only *β*_2_ CDI difference was present consistently throughout all regions.

A summary of all the proposed measures investigated and their relevance based on the seizure onset is reported in Table 2.

**Table 2.** Summary of the measures. Summary of relevant nuclei for each of our parameters. The suggested nucleus is indicated based on its prevalence of relevance throughout the analysis. A strike-through is present is a positive/negative CDI is significantly present in non-responders vs responders (i.e., not desirable), as reported in Figure 6. The correlation to risk is reported as significant as reported in Figure 4 and Supplementary Figure 4. In the ENI, we report the two nuclei with the highest ENI for that region.

| Region of Seizure Onset | Spread Functional Connectivity | Positive CDI | Negative CDI | ENI (two nuclei with highest ENI) | Correlation to Seizure Risk | Nucleus of Choice |
| --- | --- | --- | --- | --- | --- | --- |
| Cingulate | CM, MD | CM | ~~ANT~~ | CM, MD | CM, ANT, PLV | CM, MD |
| Frontal | CM, MD |  | MD | MD, ANT | MD, CM | MD |
| Insula | CM, MD | CM |  | CM, MD | CM | CM, MD |
| Lateral Temporal | PLV | ~~PLV, MD~~ | ANT | ANT, MD | ANT, CM, MD, PLV | MD, PLV |
| Mesial Temporal | ANT, PLV | ANT, PLV | CM | MD, ANT | CM, ANT, PLV, MD | ANT |
| Occipital | CM, PLV, MD | PLV |  | CM, PLV | CM, PLV | PLV |
| Parietal | CM | PLV, CM |  | ANT, PLV | ANT, PLV, MD, CM | PLV, CM |
| Subcortical | CM, MD |  | CM, MD | CM, MD | CM, MD | CM, MD |

## 4 Discussion

Thalamic electrical stimulation is a promising approach for the treatment of patients with DRE. Nevertheless, this therapeutic option currently only results in a median 50-65% seizure reduction rate at 5-years post-implantation^13(p201),22^.

An essential step toward improving outcomes for patients is in optimizing the choice of the stimulation target based on the epilepsy phenotype. Current clinical approaches favor ANT for focal epilepsies and CM for generalized epilepsies^28^. Additional nuclei, such as MD and PLV, are proposed as alternative targets only based on their anatomical connectivity to seizure onset regions. Knowledge of anatomical connectivity of these nuclei stems mostly from MRI studies in healthy subjects or animal models^35,68–73^, which carry no information about how the presence of a pathological area (the seizure onset) can modify these networks.

While our findings validate previously defined anatomical connectivity profiles, they also suggest a more nuanced strategy which recognizes that connectivity is dynamic and patient specific. That is, the connectivity between nuclei and cortical and subcortical regions can be different for different patients, but most importantly different in the epileptogenic network (i.e., including the seizure onset zone) compared to the non-pathological networks, although belonging to the same brain location.

Firstly, we revealed functional connectivity profiles to different regions of the brain for the four nuclei. The ANT exhibited the strongest connectivity to mesial temporal regions, while the MD preferentially connected with frontal areas. CM, on the other hand, showed broader connectivity to different brain areas (parietal, cingulate, subcortical, and insular regions) and PLV had strong connectivity to lateral temporal and occipital areas. These findings largely match the hodological examinations that have been done previously using anatomical connectivity^9,28^, and they also align with reports that CM stimulation is more effective for generalized epilepsy^41,74–76^, highlighting its more central role within brain networks compared to the other three nuclei.

Yet, it is possible that two or more nuclei show a similar functional connectivity to a given brain region, making it challenging to pinpoint which one may serve as the best target for neuromodulation. For this reason, we also considered a new metric of connectivity that assesses the relationships between nuclei and regions inside and outside the SOZ, comparing epileptogenic and non-epileptogenic networks. This measure, the *Connectivity Divergence Index (CDI)*, indicated that there were unique combinations of nuclei-region whose connectivities varied according to whether they were (non)epileptogenic. This result has a fundamental implication in the clinical perspective: corticothalamic connectivity is impacted by the epileptogenicity of the region only for some nuclei, which suggests their involvement in the pathological network. Understanding this relationship can further our knowledge in determining the role of each thalamic nucleus in epilepsy and seizures.

We found specific nucleus-region combinations which exhibited a high CDI, specifically: ANT-mesial temporal, MD-lateral temporal, CM-insula, cingulate, parietal and PLV-occipital, parietal and lateral temporal. Our results suggest that there may be value in using the CDI to identify the nuclei that have a strong differential connectivity to epileptogenic than to non-epileptogenic regions, rather than using connectivity as the sole metric for selecting one target over another.

Importantly, we also revealed that the four nuclei exhibit specific frequency-dependent connectivity to brain areas which are more prevalent in epileptogenic networks (i.e., SOZ-networks) compared to non-epileptogenic ones: *β*_1_ for ANT, *β*_1−2_ for MD, *δ*, *θ* for PLV and broadband *δ* − *β*_2_ for CM. The importance of *β*_1_ in ANT-based networks aligns with earlier findings from our group in which we observed that *β*_1_ ANT-cortical connectivity differed between responders and non-responders to ANT DBS^77^, and ANT stimulation significantly reduced bilateral scalp connectivity in *β*_1_. This finding corroborates the idea that this differential modulation of epileptogenic and non-epileptogenic networks only occurs within specific frequencies, which should be further explored for identifying biomarkers.

In addition, thalamic spectral power and CDI both change as the risk of a seizure increases. While this is not related to predicting the exact moment of seizure onset, existing literature has demonstrated that stimulation may have differing effects depending on whether seizure risk is high or low, underscoring the relevance of this analysis^78–81^. Besides, if a nucleus’s activity is linked to seizure occurrence, it may further validate its role in pathogenesis, thereby strengthening the rationale for choosing it as implantation target.

To further emphasize the modulating role of the thalamic nuclei in different brain states, we examined two scenarios where diverging states are present and can, in principle, affect thalamic connectivity: (i) awake versus NREM and (ii) seizure-free versus seizure days. Our findings showed that nuclei-region pairings consistently presented a different CDI in awake states than in sleep states. This supports the notion that thalamic-based modulation of epileptogenic networks is affected by sleep and wake states. Evidence that the thalamus orchestrates sleep and that sleep significantly impacts seizure occurrence supports our state-specific approach, as biomarkers may differ between awake and NREM states ^64^. More investigation is needed to study the dependency of stimulation during wake/sleep states on response to modulatory treatment. When analyzing seizure-free vs seizure-days, the ANT and PLV nuclei had higher CDI values under low-risk conditions (seizure-free days) compared to high-risk conditions (seizure days), suggesting that a heightened CDI might measure a protective mechanism (i.e., strong connectivity to seizure onset) that potentially inhibits seizure generation. In contrast, CM and MD networks displayed a similar CDI in seizure-free and seizure days.

Finally, we evaluated whether our proposed measures could predict the patient’s responsiveness to neurostimulation treatment. Interestingly, the specific nucleus-region measures that we proposed in this study were able to differentiate between the two. We indeed found that our proposed measures (functional connectivity and CDI) were different between the responders and non-responders groups in patients who received CM or PLV stimulation, further supporting their use in clinical evaluation, in a region-dependent manner. While the association with the efficacy of stimulation needs to be clarified, we believe that a baseline differential role of the nucleus in the network might aid the propagation of seizure activity or halt it based on its influence on neighboring regions. We argue that the proposed measures (connectivity, CDI, and ENI) can not only help identify the target for neuromodulation but can also aid with the therapy’s optimization and tracking of the course of the treatment, especially in closed-loop approaches.

### Limitations and Further Steps

Our study has several limitations. First, it focuses solely on inter-ictal data, excluding pre-ictal and ictal states, which could provide further insights into how pathological networks evolve during seizure phases. While inter-ictal data is often underestimated in epileptogenicity, its relevance for understanding and optimizing neuromodulatory therapies is significant, as such treatments, including DBS^14^ and RNS^18^, act as chronic modulators rather than acute seizure terminators^16^. Second, while data from multiple days in the EMU was incorporated and analyzed in a large cohort of 38 patients, it remains to be elucidated how these measures change on a longer time scale, and what is their robustness and stability over time. Importantly, the study is limited by implantation planning, as electrode locations and seizure onset sites varied across nuclei-specific subcohorts, reflecting clinical practice and potentially introducing bias. Spatial sampling limitations and a sample size of 38 patients restricted the inclusion of all seizure onset pairings in each subcohort, leaving some pairings not represented (PLV-Insula, PLV-subcortical and MD-Occipital). Lastly, the lack of clinical outcome data for most patients precluded correlating the analyzed features to therapeutic outcomes across all nuclei, as the data was collected during pre-surgical evaluation and not specifically for neuromodulatory interventions.

In conclusion, this study underscores the value of interictal data in identifying signatures and measures to support target selection for neuromodulatory treatment implantation. Our findings reveal distinct connectivity patterns among thalamic nuclei with brain regions, suggesting optimal targets: ANT for mesial temporal epilepsies, PLV for posterior or lateral temporal epilepsies, CM for cingulate, parietal, insular, and subcortical epilepsies, and MD for cingulate, frontal, insular, lateral temporal, and subcortical epilepsies (Table 2). These findings provide much-needed support for understanding the role of the thalamus in epilepsy and for the advancement of personalized neuromodulation strategies.

## Acknowledgments and Funding

The authors are deeply grateful to the patients who participated in this study and gave consent for the collection of their data for the advancement of knowledge and for a better future for epilepsy patients. The funder had no role in the experimental design, analysis, manuscript preparation, or submission. All authors had complete access to data. All authors authorized the submission of the manuscript.

This work was funded by a grant awarded to R.P. and L.I. from the Swiss National Science Foundation (SNF 197766), and a grant awarded to P.S. (CDMRP FY21 Epilepsy Research Program W81XWH-22-1-0315).

## 5 Supplementary figures

**Supplementary Figure 1.**
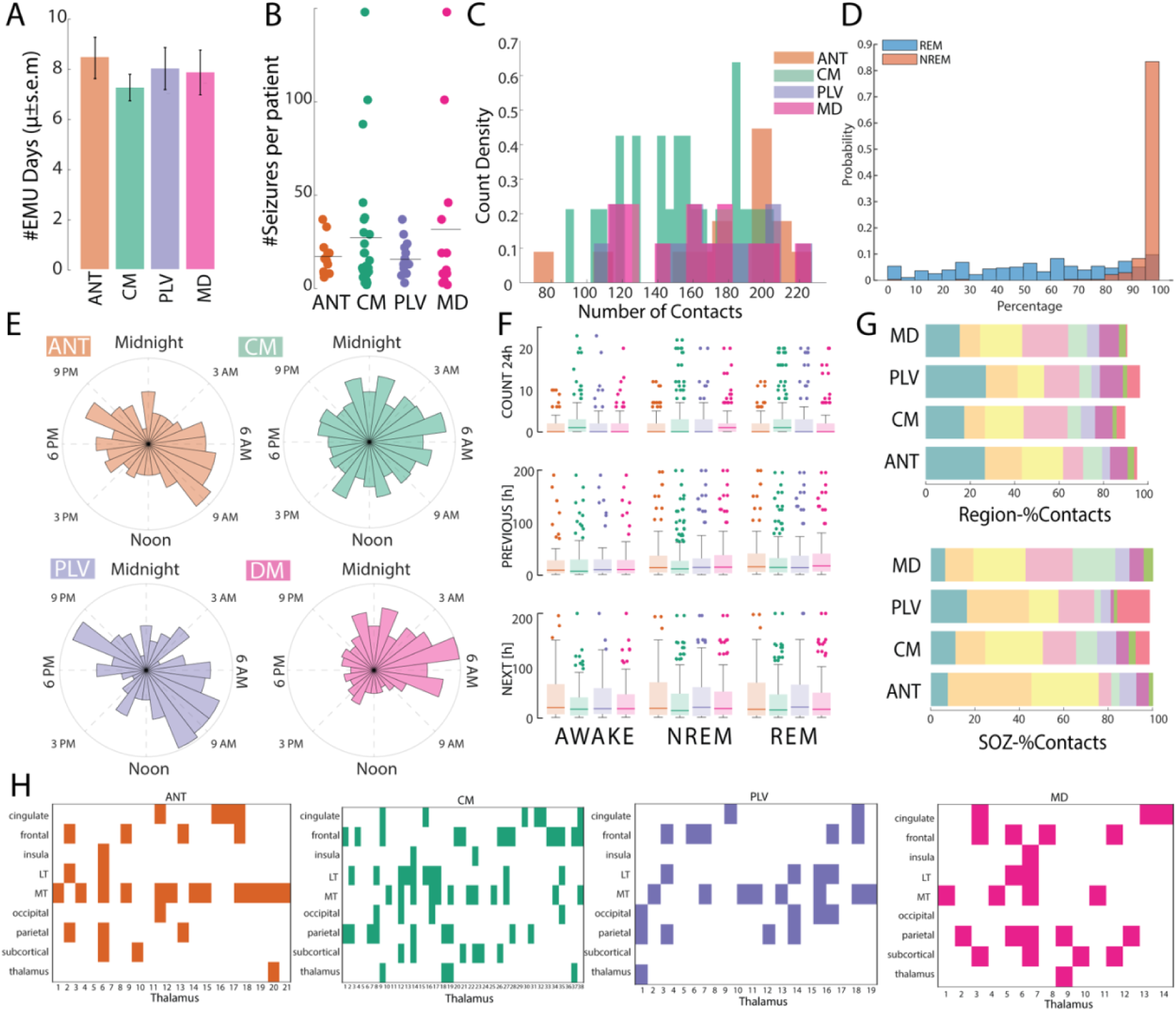
Study Design – Additional Information. **A)** Number of EMU days (*μ* ± *sem*) for each sub-cohort. **B)** Number of seizures for each sub-cohort. **C)** Number of contacts for each subcohort, across subjects. **D)** Percentage of REM/NREM in the selected periods **E**) Distribution of seizures throughout the day in the four subcohorts **F)** Number of seizures, time to next and time to previous seizure for each sub-cohort in the three states (AWAKE, NREM, REM). **G)** Percentage of contacts in each sub-region (top) and percentage of SOZ contacts in each sub-region (bottom) **H)** For each implanted thalamus, ipsilateral SOZ.

**Supplementary Figure 2.**
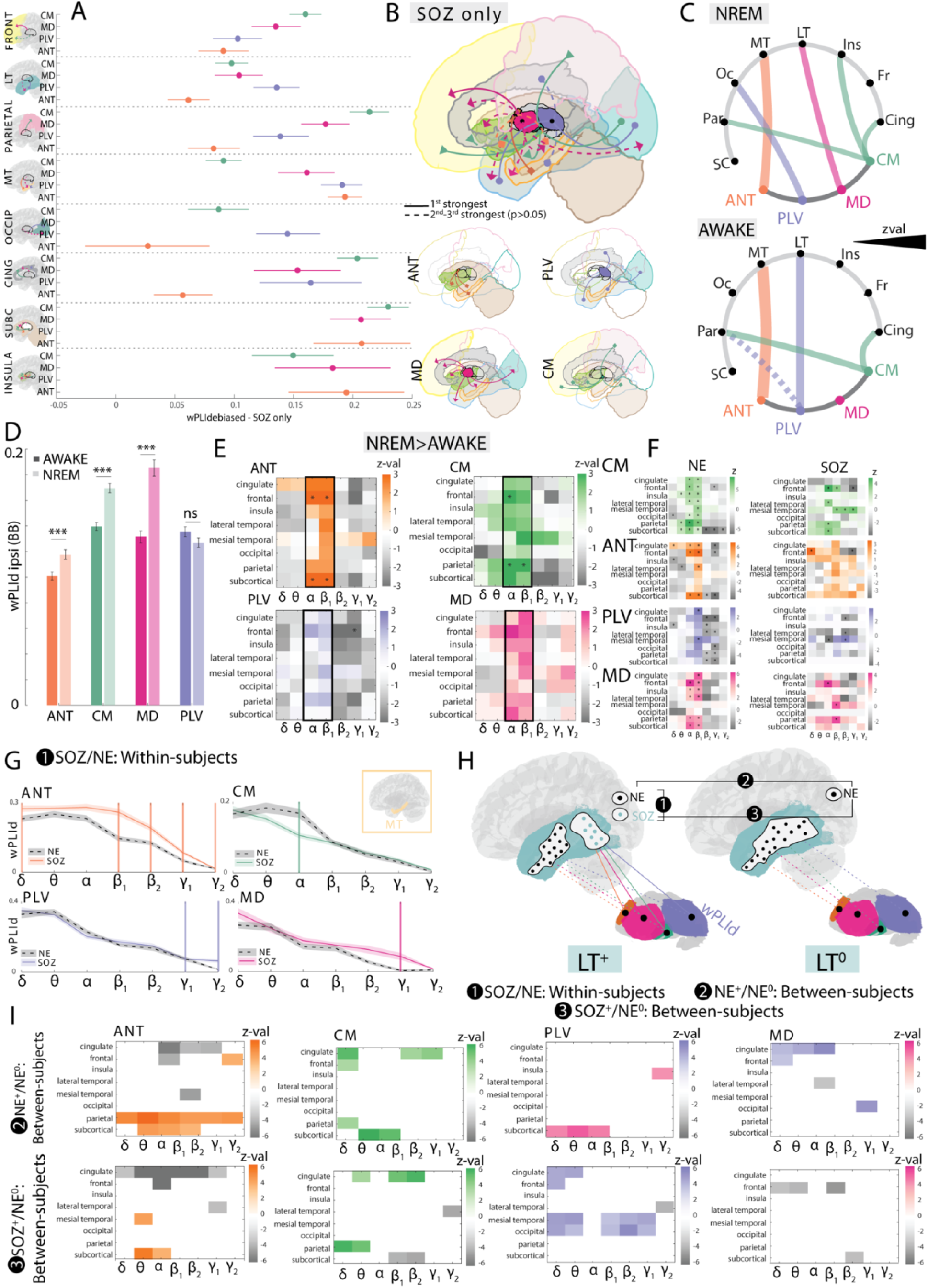
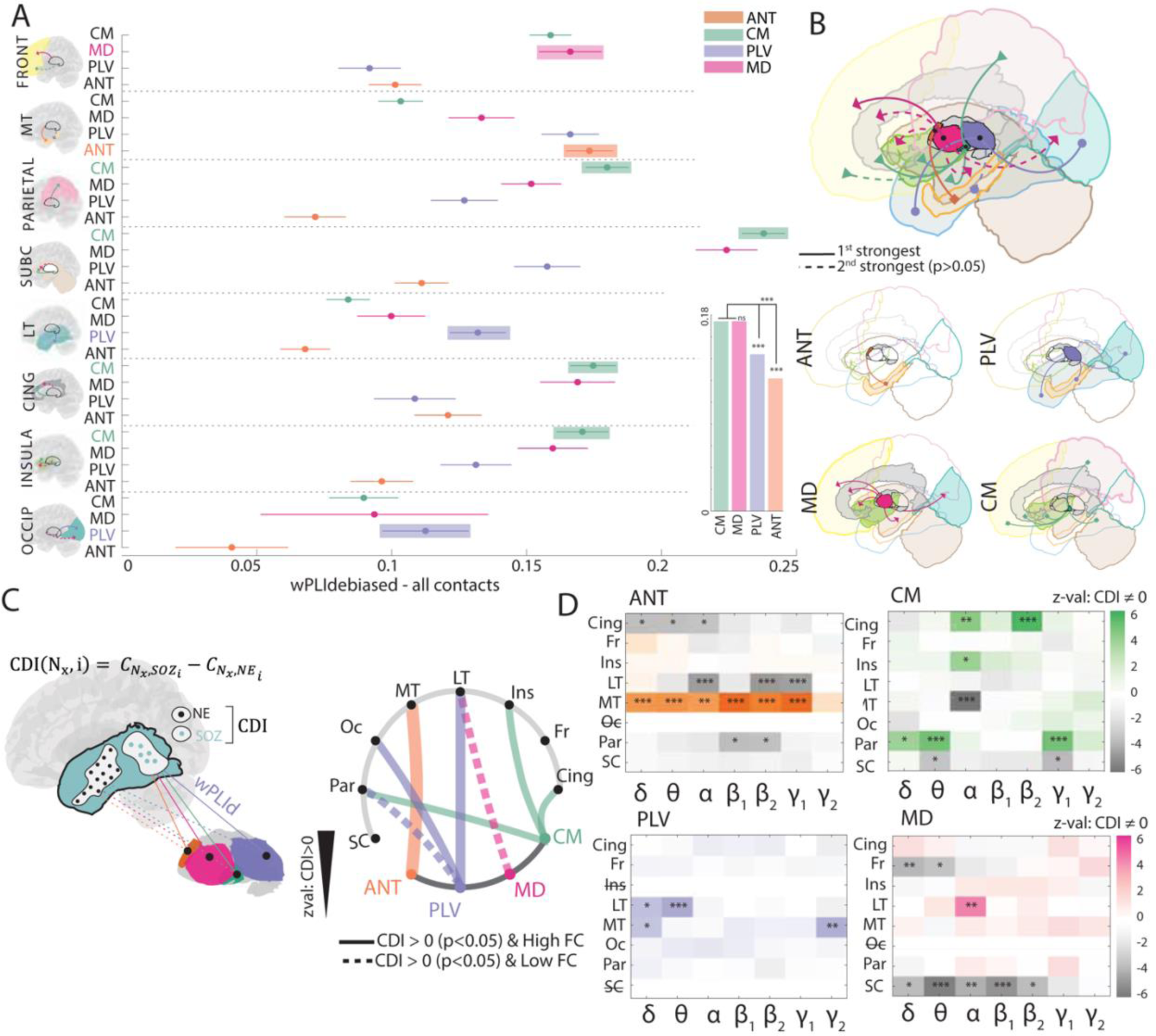
Connectivity to seizure onset and sleep/wake differences. A) Connectivity distribution for each sub-region and nucleus as reported by ANOVA-N with post-hoc Tukey-HSD (*μ* ± 95% *CI*). Connectivity is reported as broad band connectivity of each nucleus to SOZ contacts (as done in Figure 2A). B) For every region, 1^st^ (solid) and 2^nd^ (dashed, not significantly different from 1^st^) preferential connections with the nuclei are reported (top) for SOZ contacts. The same result is reported for every nucleus separately (bottom). (as done in Figure 2B) C) For each nucleus and region, we computed the CDI as the difference of nucleus connectivity to SOZ and NE separately for NREM and AWAKE. We then tested for differences using Wilcoxon signed-rank test. We report one-tailed significant comparisons (where CDI∼0, and z-val>0). The thickness of each line in the chordplot is proportional to the z-value. If the pairing was also presenting high FC to SOZ (as in panel A and B), it is represented with a solid line. Else, it is represented with a dashed line. D) Average difference in AWAKE vs NREM wPLId (Wilcoxon signed-rank) for each nucleus. E) Results for Wilcoxon signed-rank test (AWAKE vs NREM wPLId) for each region-band specific combination. F) Same result as in E but divided for SOZ and NE contacts. G) Graphical explanation of calculation of significant CDI by band. H) Graphical representation of further analyzed comparisons (unpaired). I) Results for H. If blank, result is not significant (Mann-Whitney rank-sum test).

**Supplementary Figure 3.**
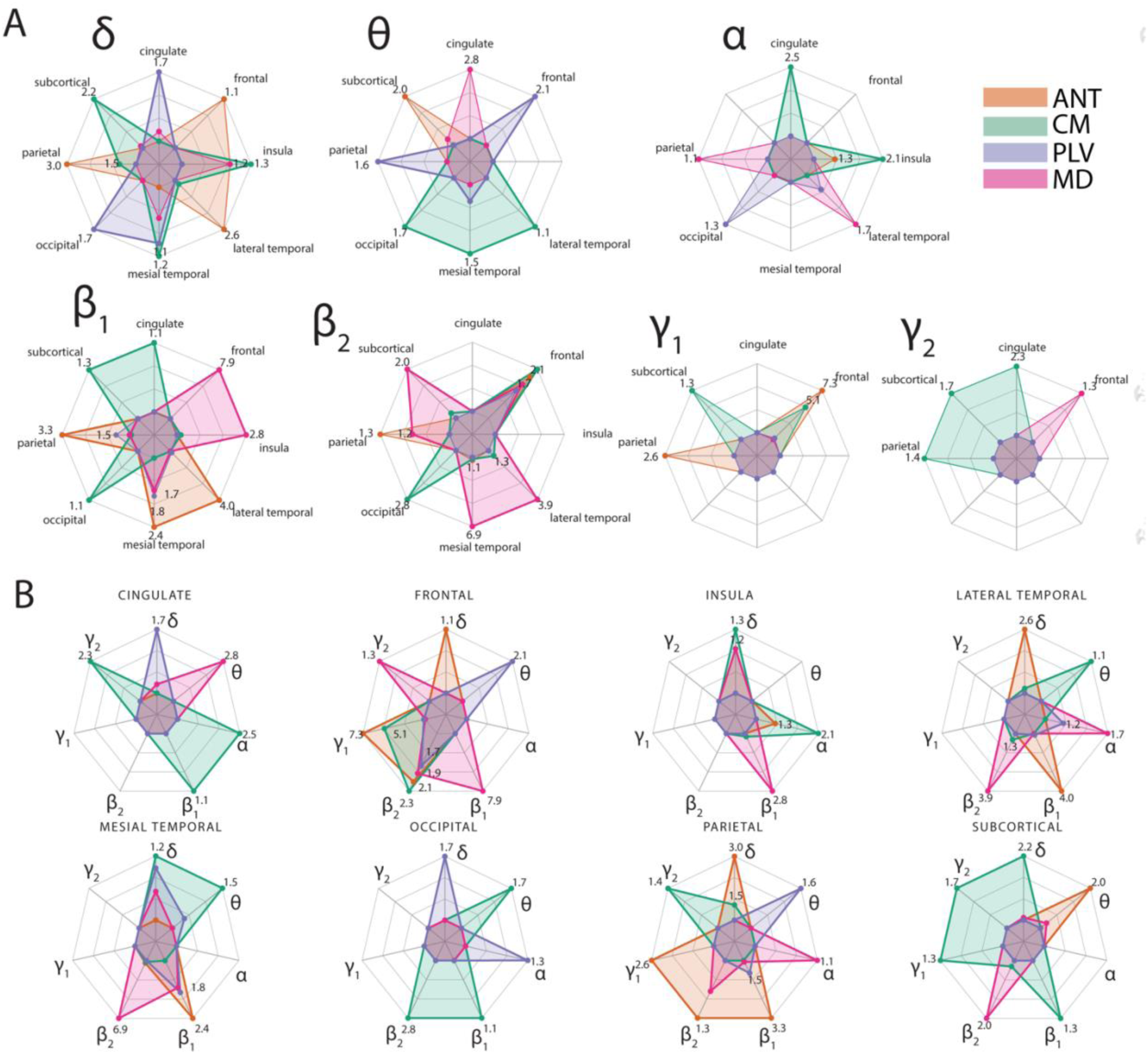
Epileptogenic-Non Epileptogenic Index: by bands and by regions. A) For every nucleus (encoded as a color) and every band, we report the ENI at every region (as in Figure 3B) B) For every nucleus (encoded as a color) and every region, we report the ENI at every band (as in Figure 3B)

**Supplementary Figure 4.**
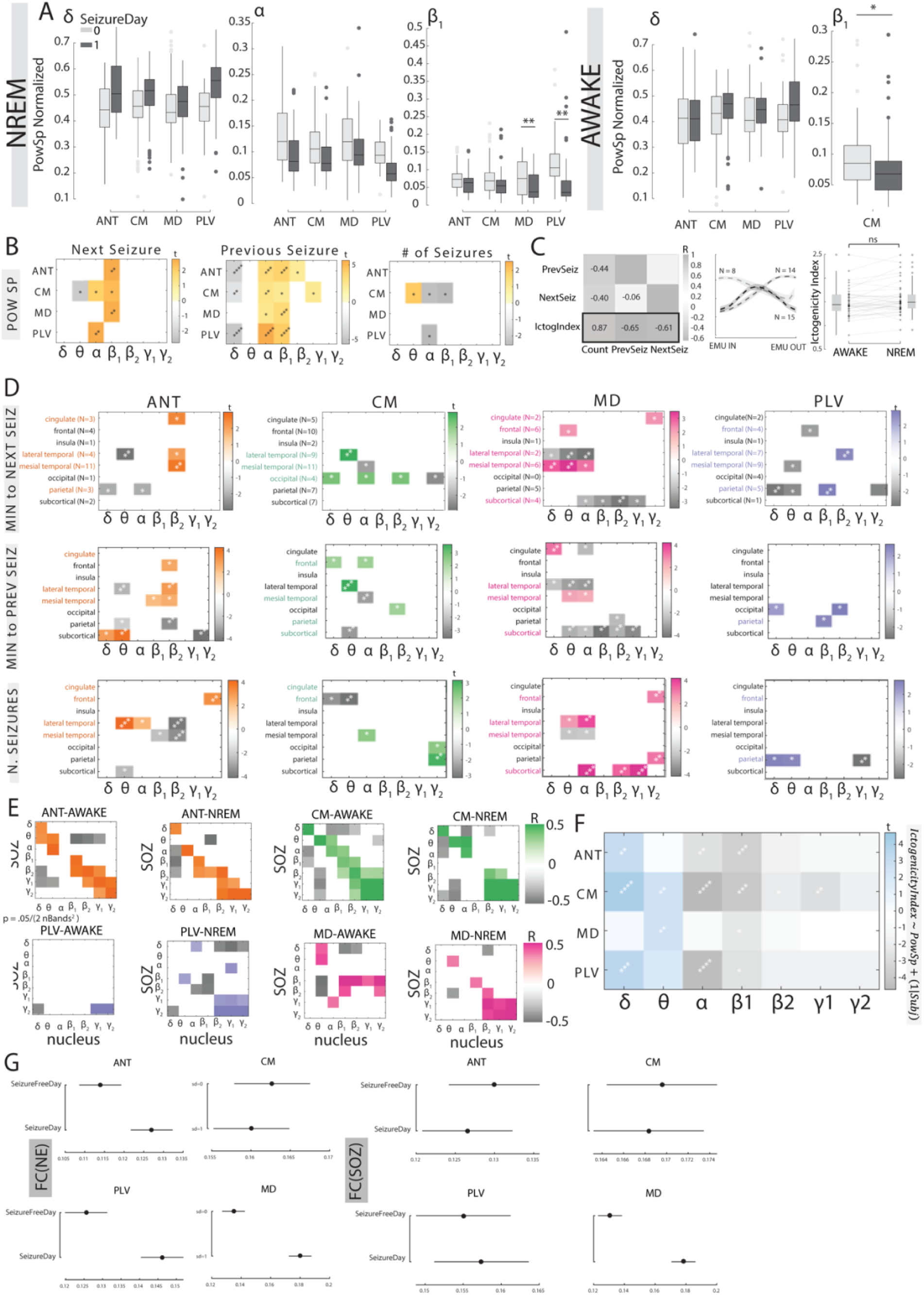
Thalamic activity correlates to number of seizures, time to next and time to previous seizures. A)Nuclei Power Spectrum in seizure-day and seizure-free days B) Thalamic power spectrum correlation (t-statistics, LMM) to ictal measures C) Correlations of ictal measures to the Epileptogenicity Index (Left), Clusters of evolution of the Epileptogenicity Index throughout the EMU stay (Middle), Differences in Epileptogenicity Index in wake/NREM states D) Correlations of CDI to the ictal measures (t-statistics, LMM) E) Correlation of thalamic power spectrum to SOZ power spectrum for the four nuclei, in NREM and wake state E) Correlation of thalamic power spectrum to the Epileptogenicity Index G) Functional connectivity to NE and SOZ areas in seizure-free and seizure-days for the four nuclei

**Supplementary Figure 5.**
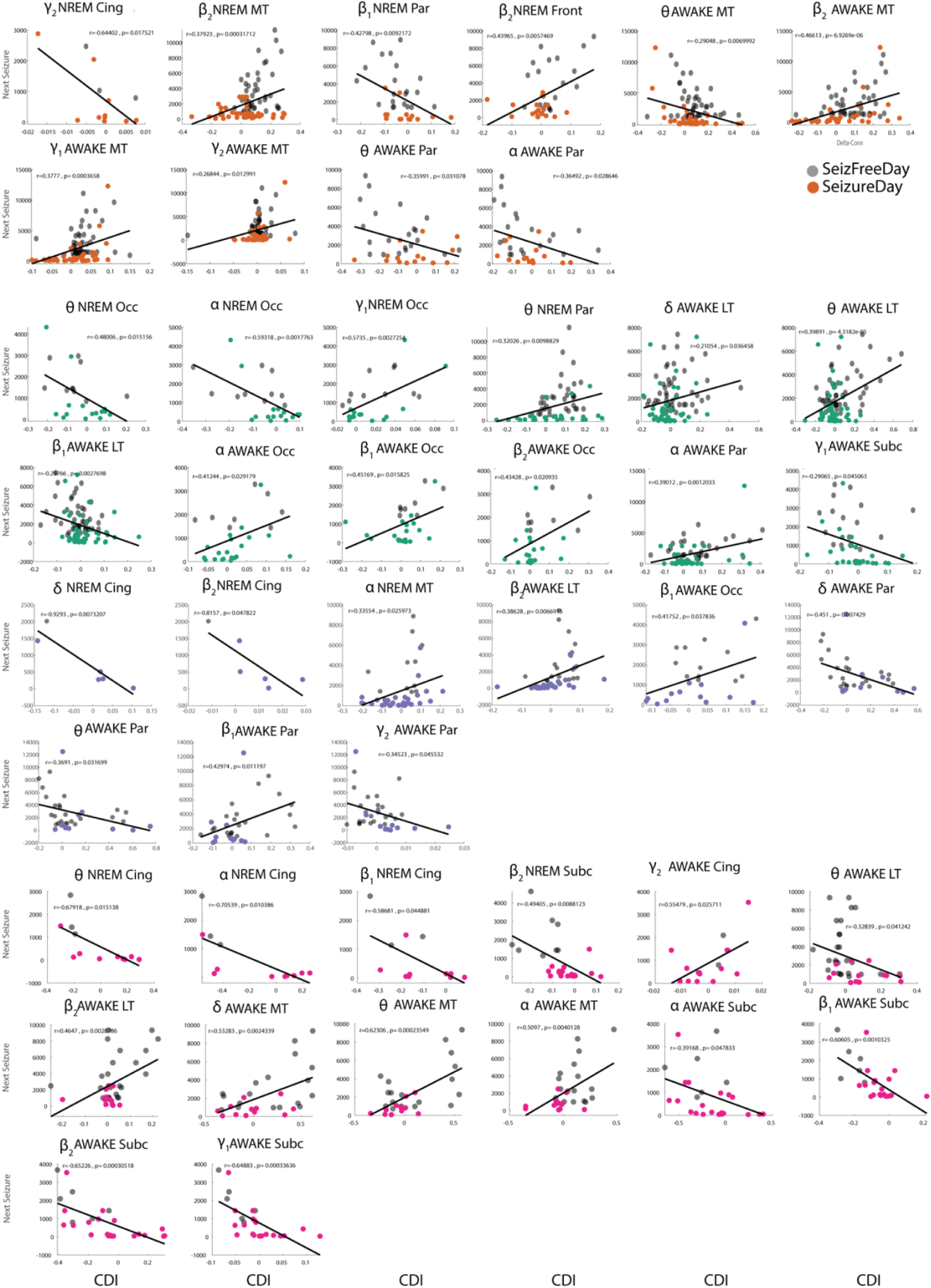
Thalamic connectivity is modulated based on the time to next seizure. Pearson’s correlation (along with Pearson’s R and p-values) between band-nucleus-region CDI and time to the next seizure. Seizure-free days are represented as colored markers (relative to each nucleus), while seizure-days are in black.

